# Longitudinal Central Adiposity Accumulation is Associated with Cortical Alteration and Impaired Cognitive Function in Adolescents

**DOI:** 10.64898/2026.04.22.26351453

**Authors:** Lingxuan Zhang, Bowen Qiu, Zhifan Chen, Xinyi Xu, Ruoke Zhao, Yiwei Chen, Chenglin Ning, Ruike Chen, Mingyang Li, Dan Wang, Junfen Fu, Dan Wu

**Author notes:** Corresponding to: Junfen Fu, PhD and Dan Wu, PhD, and.

## Abstract

Childhood obesity remains a pressing global health challenge, yet the impact of dynamic adiposity changes during active developmental window retains poorly understood. Leveraging longitudinal data from the Adolescent Brain Cognitive Development (ABCD) Study (N=8519 at baseline; N=1873 at 4-year follow-up), our study reveals distinct neurodevelopmental implications of central fat dynamics during adolescence. At baseline, central fat indices (body roundness index, BRI / waist-to-height ratio, WHtR) outperformed BMI in predicting cognitive deficits, showing robust associations with impaired inhibitory control and episodic memory. The prediction effect was partially mediated by cortical changes in prefrontal and temporal regions. Longitudinally, the rate of fat accumulation (Δ) emerged as a critical predictor: faster adiposity accrual predicted attenuated cortical thinning (i.e., slower development) in parietal lobes and poorer executive function at follow-up, while baseline adiposity showed no significant effects on the follow-up brain morphology or cognitive development. Notably, subgroup analyses uncovered that obese adolescents with central fat reduction exhibited accelerated cortical thinning in posterior cingulate (change difference p=0.006-0.029) alongside rapid improvement in inhibitory control (Flanker slope difference p<0.05), whereas those with persistent adiposity showed delayed thinning in the postcentral gyrus. The study reveals that central fat (BRI/WHtR) is closely linked to neurocognitive risks, and longitudinal fat accumulation—rather than baseline adiposity—drives cortical alteration. Notably, fat reduction activated adaptive neural change in obese adolescents, underscoring the importance of weigh regulation during neurodevelopment.

## Introduction

Childhood obesity remains an urgent global public health issue. According to the World Health Organization (WHO), over 18% of children and adolescents aged 5 to 19 years are either overweight or obese, who carry high probability of persistence into adulthood(Sahoo et al., 2015). Obesity not only increases risks of metabolic disorders(El Meouchy et al., 2022; Ng et al., 2021; Pulgaron and Delamater, 2014), but also is associated with impairments in cognitions, including executive function, attention, visuospatial processing, and motor skills(Liang et al., 2014; Likhitweerawong et al., 2022; Nazmus Sakib et al., 2023). Despite extensive neuroimaging research on obesity in adults, studies focusing on children remain relatively scarce. Childhood obesity may involve distinct neurodevelopmental mechanisms compared to adulthood, given the rapid brain development, heightened neuroplasticity, and unique hormonal and metabolic profiles during this period. While cross-sectional studies in children have provided valuable insights, they cannot capture developmental changes over time, underscoring the need for longitudinal research to clarify how obesity-related brain alterations emerge and evolve during growth.

Indeed, existing cross-sectional neuroimaging studies have revealed reduced cortical thickness and volume in obese children(García-García et al., 2019; Laurent et al., 2020; Wierenga et al., 2014), particularly in regions governing cognitive control and emotional regulation (e.g., the prefrontal cortex, cingulate cortex, hippocampus)(Kaltenhauser et al., 2023). These alterations might mediate the association between adiposity and cognitive dysfunction (Nazmus Sakib et al., 2023; Ronan et al., 2020). Given that body mass index (BMI; calculated as weight in kilograms divided by height) inadequately characterizes body composition (Khan et al., 2023), recent research increasingly incorporated measures that better capture central fat distribution, such as body roundness index (BRI) (Thomas et al., 2013) and waist-to-height ratio (WHtR), which have been shown to exhibit stronger associations with obesity-related metabolic diseases(Chen et al., 2024; Rico-Martín et al., 2020; Zhang et al., 2024). Importantly, both BMI and central fat indices have been associated with cortical morphology, but reported effects were often age-dependent, e.g., greater visceral fat was found to be associated with a smaller gray matter volume in 9-10 year-old children (Silva et al., 2021), whereas other studies reported higher cortical thickness in adolescents aged 15–18 years (Saute et al., 2018; Westwater et al., 2019), in contrast with the cortical thinning and volume reduction in adults aged 19–50 (Veit et al., 2014; Westwater et al., 2019).These heterogeneous findings highlight the complexity of adiposity-brain relationships and raise the possibility that the fat accumulation, rather than adiposity at single timepoint, may be a key determinant of neurodevelopmental outcomes.

However, existing studies mostly focus on cross-sectional analysis or examine the impact of obesity at a single timepoint, without accounting for the dynamic accumulation of adiposity over time. This gap is critical because childhood and adolescence are sensitive neurodevelopmental windows characterized by rapid and non-linear changes in both body composition and cortical structure. In order to understand the complex and dynamic relation, longitudinal studies are ideal. A UK Biobank study demonstrated adverse effects of midlife adiposity accrual(Zhang et al., 2025) in a population of 47.6-62.8 year old adults. Kaltenhauser et al. found the impact of adiposity on neurodevelopment, indicating that higher baseline adiposity is associated with altered neurodevelopmental trajectory, attenuating typical adolescent cortical thinning (Kaltenhauser et al., 2023). Chiesa et.al demonstrated that childhood adiposity contributes to adult brain traits such as smaller grey matter volume (Chiesa et al., 2025). Yet, the impact of dynamic adiposity accrual velocity during adolescence remains uncharacterized, partially due to the lack of data in this period. It would be intriguing to know how the speed and trajectory of weight gain during this sensitive neurodevelopmental window influence cortical maturation.

This study leveraged longitudinal data from the Adolescent Brain Cognitive Development (ABCD) Study to investigate the impact of adiposity on the adolescent brain, with two primary aims: (1) To determine the role of longitudinal adiposity accumulation during this sensitive neurodevelopmental period on brain development and cognitive outcomes. (2) To compare the predictive power of generalized adiposity (measured by BMI) versus central adiposity (measured by WC, BRI and WHtR) on neurocognitive outcomes. We hypothesized that the adiposity accrual rate, particularly of central fat accrual, had more strongly influences on brain development and cognitive outcomes, compared to static status at single visit. The analytical pipeline is outlined in Fig. 1.

**Figure 1.**
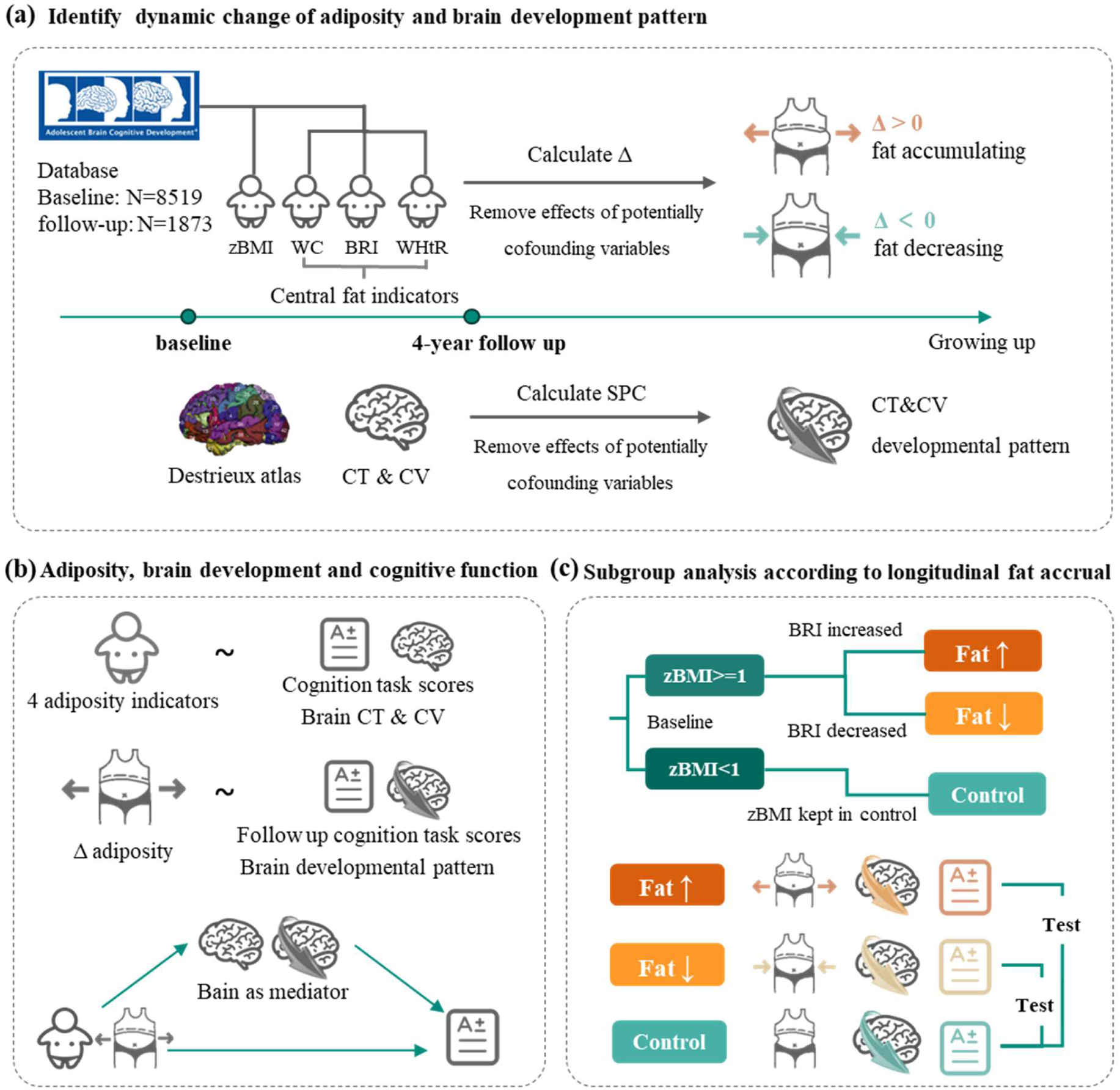
The study workflow. The analysis pipeline incorporated three sequential components. (a) Calculation of adiposity accrual velocity using four anthropometric indicators (zBMI, WC, BRI, WHtR) and brain developmental patterns via symmetrical percentage change (SPC) from longitudinal neuroimaging. (b) A two-stage analytical approach employing linear mixed-effects models to examine both cross-sectional associations (baseline adiposity metrics vs. cognition/brain morphology) and longitudinal relationships (adiposity accrual velocity vs. follow-up outcomes), with mediation analysis of neural pathways. (3) Subgroup analysis of obese children between those showing fat accumulat versus reduction, in terms of neurocognitive trajectories, cognitive and neurodevelopmental outcomes. respectively. Abbreviation: zBMI, body mass index z-score, calculated according to the WHO Child Growth Standards; WC, waist circumference; BRI, body roundness index; WHtR, waist-to-height ratio; Δ, rate of change, calculated as the difference in indicators divided by the difference in age; CT, cortical thickness; CV, cortical volume.

## Methods

### Participants and dataset

This study utilized prospective cohort data from the Adolescent Brain Cognitive Development (ABCD) study, a longitudinal population study of brain development in young adolescent between 9-11 years old (https://abcdstudy.org). This investigation used the baselines and 4-year follow-up data from ABCD Data Release 5.0, and further excluded participants with (1) a history of anti-obesity medication use (n=32), (2) extreme adiposity indicators values (n=483), or (3) missing data for neuroimaging data or key covariates (n=455) (details in the Supplementary Methods). Comprehensive written consent from parents and enthusiastic assent were obtained from all participants. Rigorous research protocols were reviewed and approved by the institutional review board of all participation sites.

### Adiposity measures

In the ABCD protocol, participants’ height and weight underwent triplicate measurements with averaged values (https://nda.nih.gov/data-structure/abcd_ant01). The body mass index z-score(zBMI) was calculated according to the WHO Child Growth Standards(de Onis et al., 2007). WC was measured around the abdomen at the level of the iliac crest level (https://nda.nih.gov/data-structure/abcd_ant01). BRI was calculated as 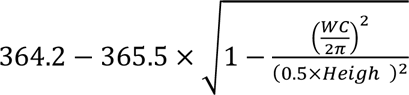 (Thomas et al., 2013), and WHtR was calculated as 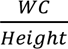. The adiposity velocity accrual rate was calculated as 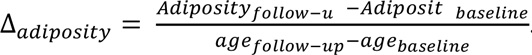.

### Cognitive Function

This study utilized standardized neurobehavioral assessments based on the National Institutes of Health (NIH) Toolbox provides (Weintraub et al., 2013), comprising 7 validated measures of executive function, working memory, processing speed, attention, episodic memory, and language abilities (Luciana et al., 2018; Weintraub et al., 2013). The battery is applicable across a broad age range from 3 years onward. Due to the high incomplete data in two of the cognitive tasks (list sorting working memory=0.603, dimensional change card set=0.440) at 4-year follow-up, this investigation used the following 5 cognitive tasks: the Flanker task (inhibitory control), the pattern comparison processing speed test (speed of visual information processing), the picture sequence memory test (episodic memory), the picture vocabulary task (vocabulary) and the oral reading recognition task (phonologic processing). Higher scores on these tasks indicate better performance. Symmetrical percentage change (SPC) of each cognitive task score was calculated to represent the cognitive development.

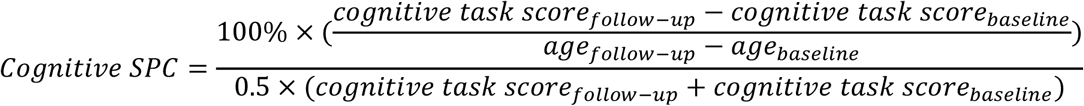

### Neuroimage data

In the ABCD study, 3D T1- and T2-weighted structural images were obtained using 3T scanners across 21 data collection sites. The scanning and preprocessing details have been previously described (Casey et al., 2018). This study used the preprocessed data including cortical volume (CV) and cortical thickness (CT), segmented based on the Destrieux cortical atlas with 148 regions of interest (ROIs) (Destrieux et al., 2010). SPC of each region of CT and CV was calculated to represent the cortical developmental pattern.

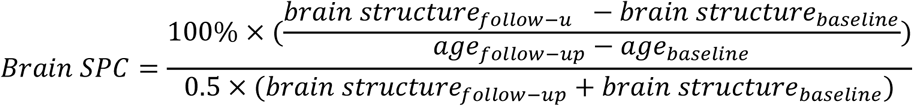

### Statistical Analysis

Statistical analyses were performed using R version 4.3.1 (R Project for Statistical Computing). This investigation included linear-mixed-effects (LME) analysis using the lme4 R package (Bates et al., 2015) and mediation analysis using the Mediation R package(Tingley et al., 2014). All reported P-values underwent false discovery rate (FDR) correction, with an adjusted P-value < 0.05 considered statistically significant.

Cross-sectional associations between adiposity indicators (zBMI, WC, BRI, WHtR) and cognitive task scores were estimated using LME models, adjusting for the covariates of sex, age, race, puberty developmental category, family income, parental education level and site. Lifestyle factors were also included as covariates, comprising physical activity (moderate-to-vigorous physical activity, muscle-strengthening activity and school-based activity) and dietary habits (quantified using parent-reported nutritional assessment scores (Morris et al., 2015)).

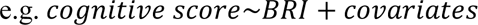

Subsequent models evaluated the independent contribution of central fat indicators (WC, BRI, WHtR) to cognition by simultaneously entering each central indicator along with zBMI into the model to examining the effect of central fat independent of overall adiposity:

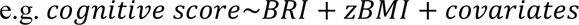

Similarly, LME analyse was also performed to investigate associations between adiposity indicators and brain morphology, incorporating the same covariates. For analyses involving CV, ICV was additionally included as a covariate. Mediation analysis was then applied to examine potential mediating effects of adiposity-associated brain morphological alterations on the relationship between adiposity and cognitive function. For each significant brain region identified in the morphology-adiposity analysis, the average causal mediation effect (ACME) was used to represent the mediating effect, followed by FDR correction.

Longitudinal analysis utilized LME analysis to estimate the relationship between Δadiposity and dependent variables (cognitive task scores and brain SPC), adjusting for baseline values in addition to the covariates used in the cross-sectional analyses:

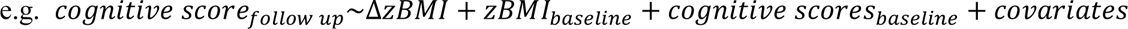

Mediation models, analogous to the cross-sectional approach, were then applied to examine whether cortical developmental trajectories mediated the association between Δadiposity and follow-up cognitive outcomes.

Furthermore, differences in cognitive and brain developmental variables between obese subgroups (increasing/decreasing BRI, classified based on whether ΔBRI ≥ 0) and the control group were assessed using the T-test for normally distributed continuous variables and the Wilcoxon rank-sum test for non-normally distributed or ordinal variables.

## Results

### Participants’ Demographic Characteristics

A total of 8519 children (mean±SD, 9.90 ± 0.62 years) with complete data at baseline were included in the baseline cross-sectional analysis. Of those, 1873 children (14.10 ± 0.69 years) had complete data at the 4-year follow-up visit and were included in the longitudinal analyses. Baseline and follow-up characteristics of ABCD participants are presented in Table 1.

**Table 1.**
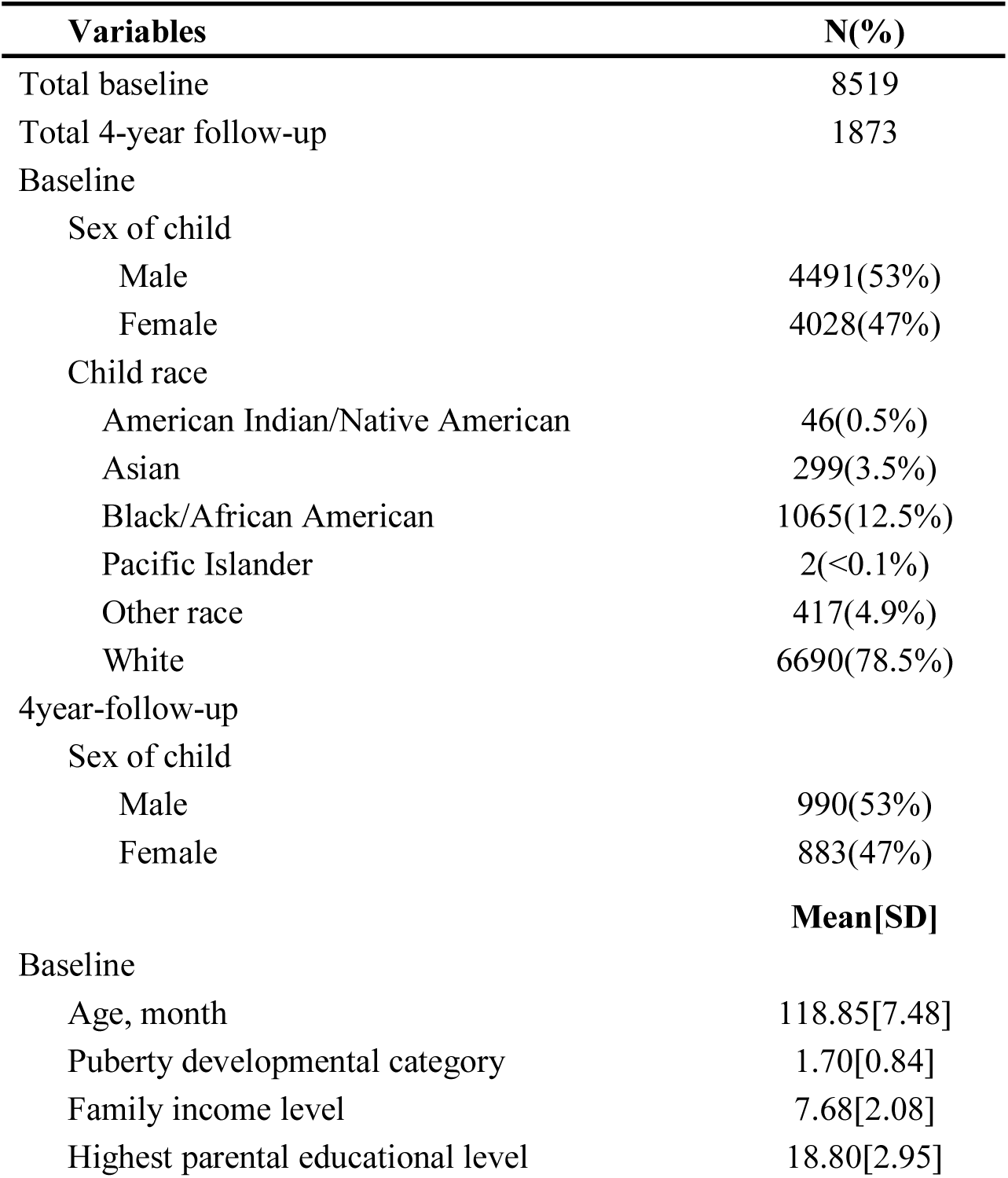
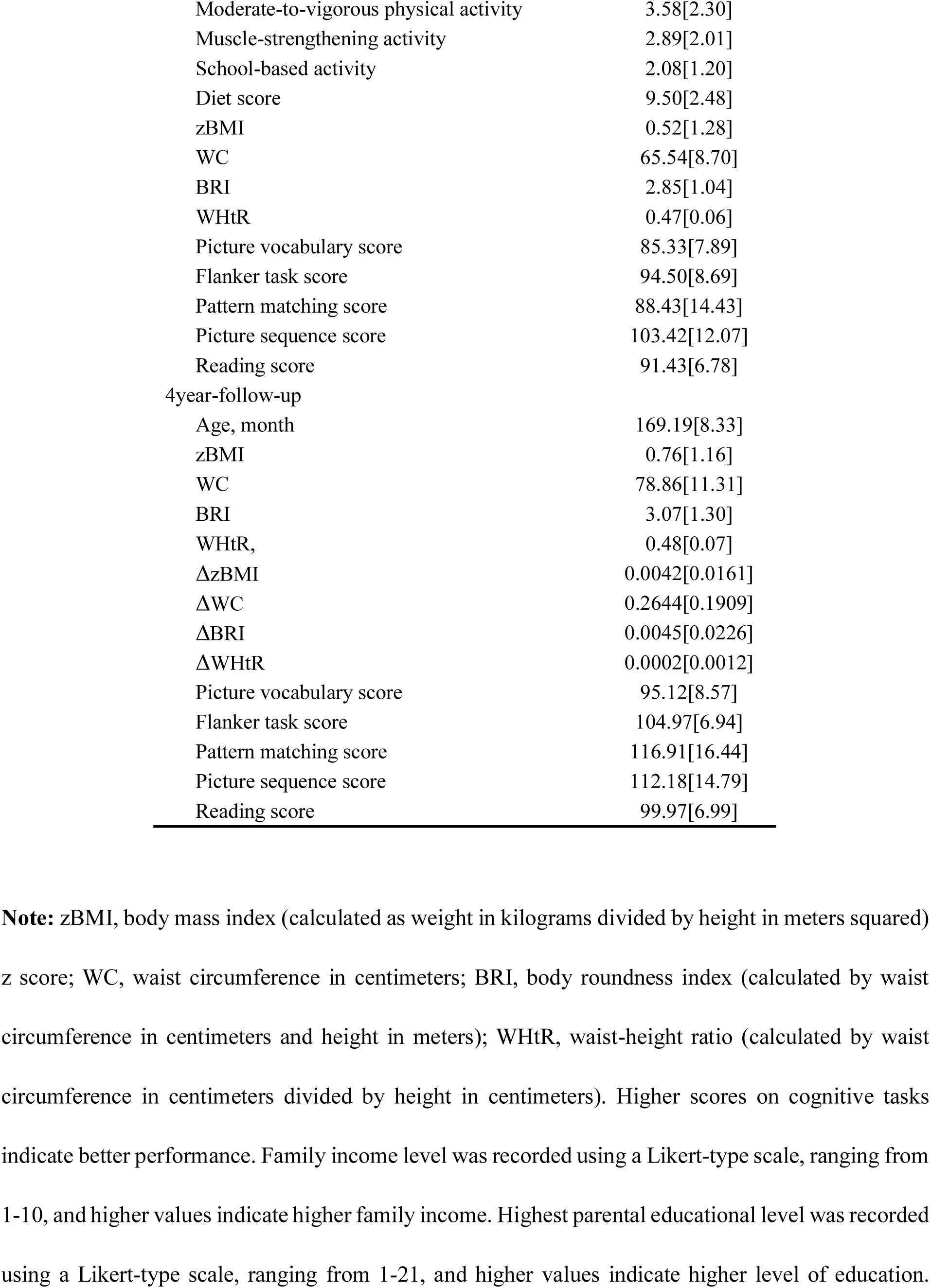
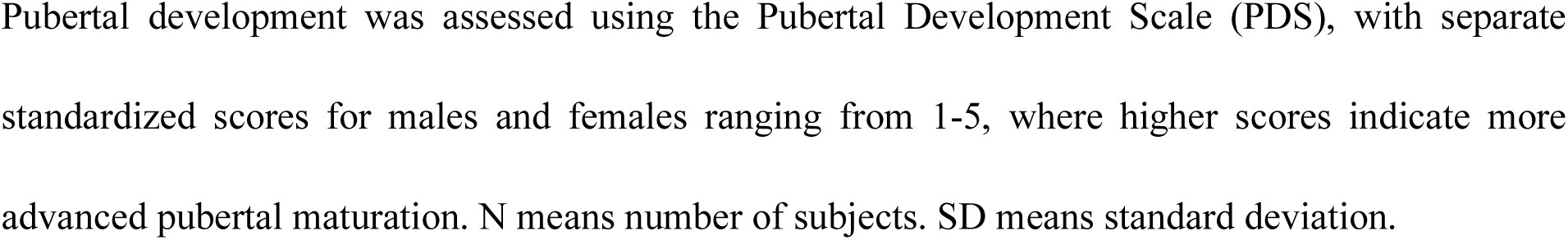
The demographic, adiposity and lifestyle statistics of the study population.

### Cross-sectional Association between Adiposity, Cognition and Cortical Morphology

Overall, higher zBMI, WC, BRI and WHtR were associated with lower cognitive task scores (Fig. 2a, Table 2). Specifically, higher BRI and WHtR were both associated with lower flanker task score (BRI: β = −0.28, adjusted p = 0.005; WHtR: β = −5.16, adjusted p = 0.006). Higher zBMI, WC, BRI and WHtR were associated with lower picture sequence score (zBMI: β = −0.42, adjusted p < 0.001; WC: β = −0.04, adjusted p = 0.027; BRI: β = −0.42, adjusted p = 0.005; WHtR: β = −7.51, adjusted p = 0.006). Higher zBMI, BRI and WHtR were associated with reading score (zBMI: β = −0.16, adjusted p = 0.010; BRI: β = −0.19, adjusted p = 0.010; WHtR: β = −3.27, adjusted p = 0.015). Moreover, the joint model of zBMI and central fat indicators (WC, BRI and WHtR) showed that the significant negative association between BRI/WHtR and flanker task score remained (BRI joint model: β = −0.45, adjusted p = 0.004; WHtR joint model: β = −8.28, adjusted p = 0.005), while zBMI was not statistically significant in these models (Table S1), indicating that BRI and WHtR were more indicative of cognitive outcomes compared to BMI.

**Figure 2.**
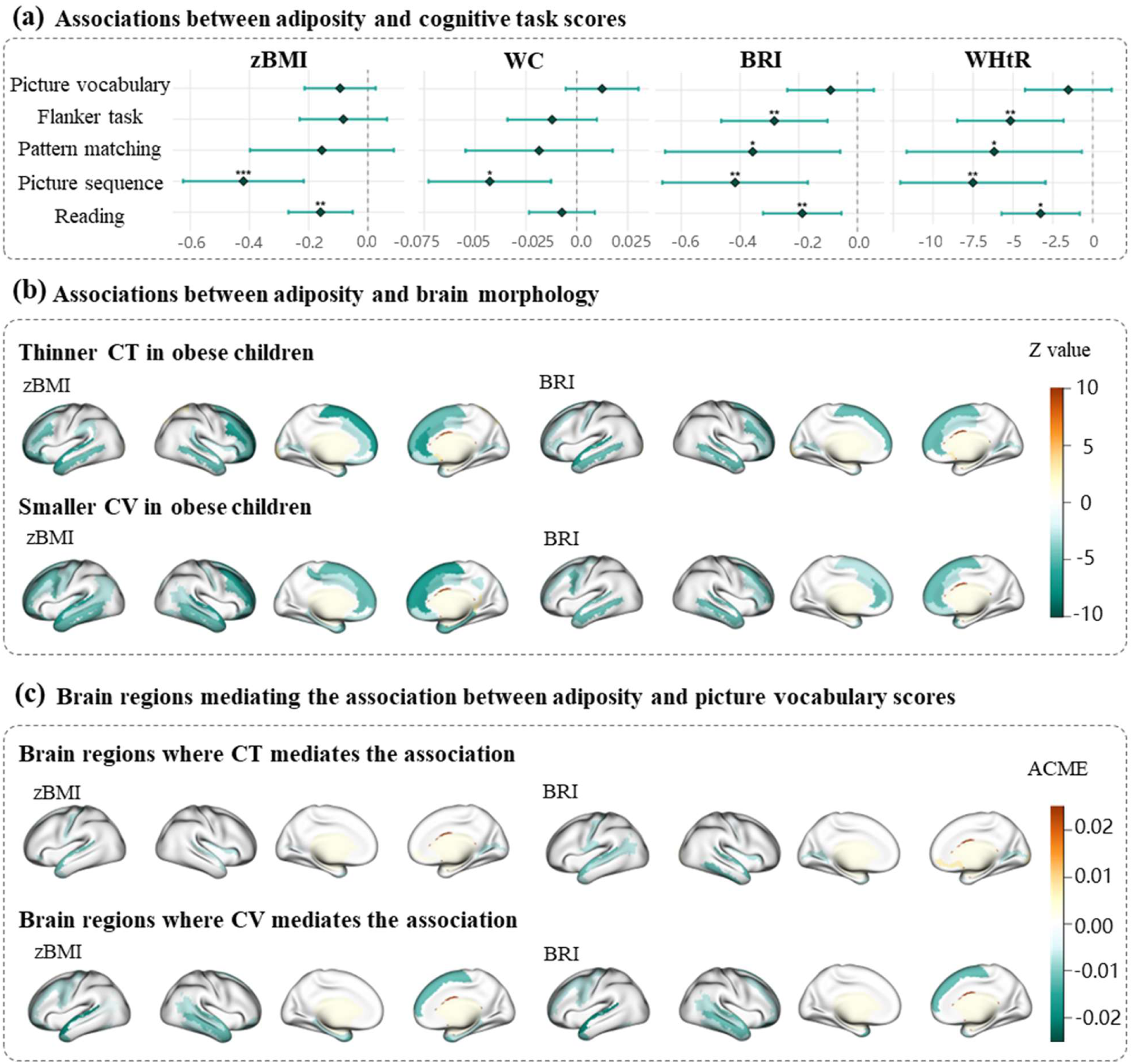
Associations between adiposity indicators, and neuroimaging features, and neurocognitive characteristics. (a) The associations between adiposity and cognitive task scores. * p<0.05, **p<0.01, ***p<0.001 (FDR-adjusted). (b) The associations between adiposity and brain morphology. The colored brain areas represented the significant associations after FDR correction, and the value indicated the z-value. (c) Significant mediating brain regions for a representative cognitive task (picture vocabulary score). The colored brain areas represented the significant mediation effects, and the value indicated the average casual mediation effects (ACME). Abbreviation: BMI, body mass index; WC, waist circumference; BRI, body roundness index; WHtR, waist-to-height ratio; CT, cortical thickness; CV, cortical volume.

**Table 2.**
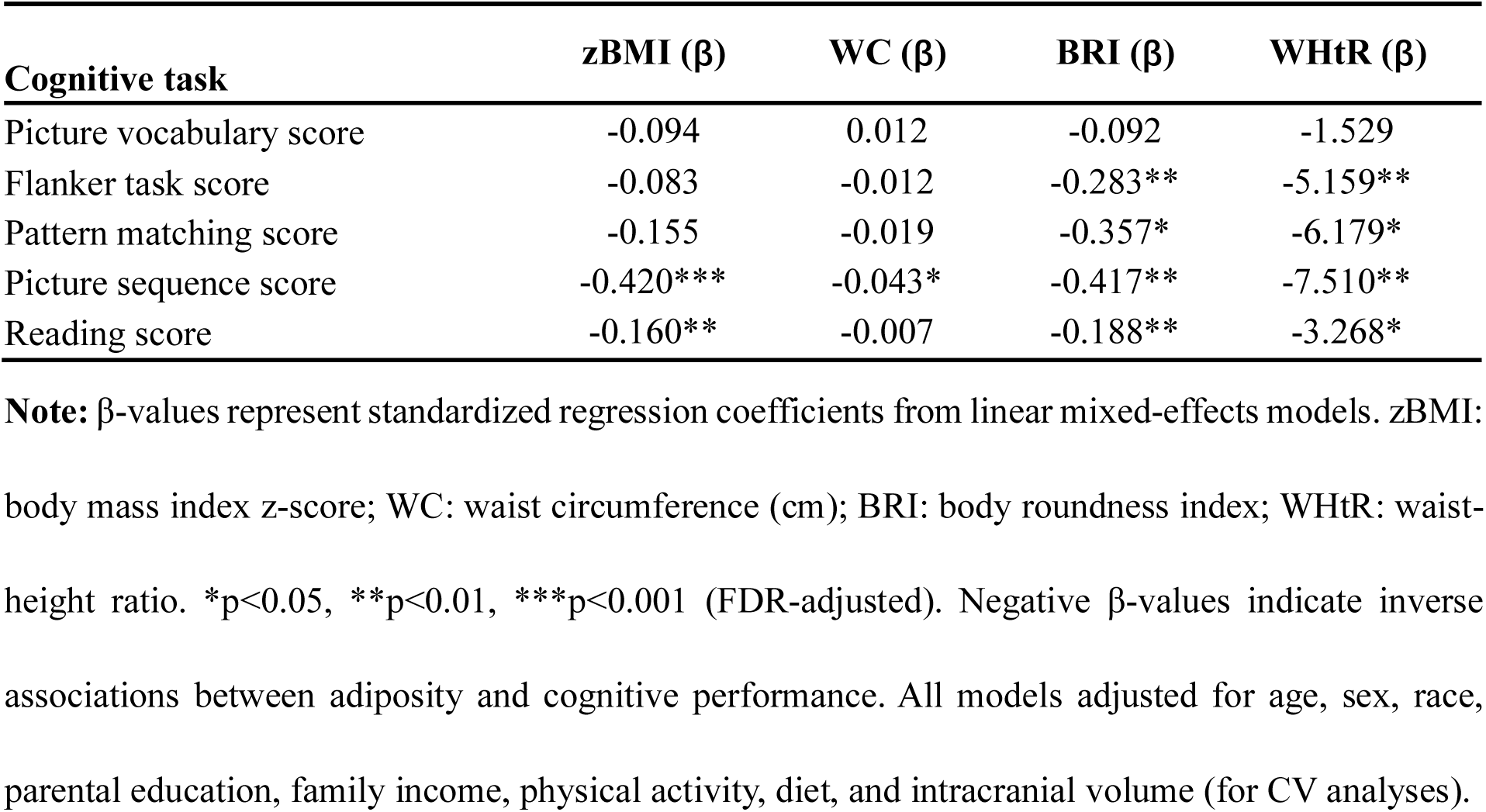
Association between adiposity indicators and cognitive task scores.

As for brain morphology, higher adiposity indicators were predominantly associated with thinner CT and smaller CV in both hemispheres (Fig. 2b, Fig.S1). Specifically, the strong correlations were concentrated in the frontal and temporal cortical regions between CT/CV and zBMI/WC/BRI/WHtR (e.g., CT of left superior temporal gyrus: β = −0.29 to −0.00, adjusted p <0.001; CV of left middle frontal gyrus: β = −1193.6 to −11.08, adjusted p <0.001).

Global mean CT and global CV were significant mediators between all adiposity indicators and picture vocabulary score as well as reading score. Regionally, CT in 4-22 regions of the 148 regions and CV in 12-26 regions partially mediated these adiposity-cognition relationships (Fig.S2-5). Notably, the anatomical locations of these mediating regions (primarily in frontal and temporal cortices) correspond to key nodes of functional networks involved in somatomotor processing (somatomoter network, SMN), internal thought (default mode network, DMN), cognitive control (frontoparietal network, FPN), and emotion (limbic network, LM) (Yeo et al., 2011), potentially reflecting a distributed functional impact of adiposity on cognition.

For flanker task score, its relationships with central adiposity (BRI and WHtR) were significantly mediated by global CV, and regionally, by 2-8 specific brain regions. As for pattern matching score and picture sequence score, while global mean CT and global CV were not significant mediators, CT in 0-11 regions and CV in 1-7 regions partially mediated these relations (Fig. 2c, FigS.2-5). These significant regions were concentrated in the temporal cortical regions, including the superior temporal gyrus (ACME = −0.47 to −0.00, adjusted p < 0.05), middle temporal gyrus (ACME = −0.47 to −0.00, adjusted p < 0.05), inferior temporal gyrus (ACME = −0.25 to −0.00, adjusted p <0.05), and temporal pole (ACME = −0.21 to −0.00, adjusted p < 0.05).

### Longitudinal Association of Adiposity Accrual with Cognitive Decline and Brain Development

Our longitudinal analysis revealed distinct patterns in the relationship between adiposity accrual and cognitive function. When examining both static adiposity measures (at baseline and 4-year follow-up) and their changes over time, we observed that the rate of adiposity change, rather than single measurement at baseline or follow-up, showed significant associations with subsequent cognitive performance (Table 3, Table S2). Follow-up flanker task score was significantly associated with ΔWC (β = −3.26, adjusted p = 0.006), ΔBRI (β = −20.21, adjusted p = 0.049) and ΔWHtR (β = −351.81, adjusted p = 0.049). Similar patterns were observed for pattern matching score (ΔWC: β = −5.82, adjusted p = 0.029; ΔBRI: β = −43.20, adjusted p = 0.049; ΔWHtR: β = −780.20, adjusted p = 0.049), emphasizing the role of central adiposity on cognitive development.

**Table 3:** Differential contributions of 4-year follow-up adiposity and adiposity accrual rate on follow-up cognitive performance in joint models.

| Adiposity | Cognitive task | 4year ($\beta$) | $\Delta$ ($\beta$) |
| --- | --- | --- | --- |
| zBMI | Picture vocabulary score | -0.165 | -4.259 |
|  | Flanker task score | -0.320 | -6.957 |
|  | Pattern matching score | -0.125 | -48.404 |
|  | Picture sequence score | -0.515 | 12.894 |
|  | Reading score | -0.046 | -5.750 |
| WC | Picture vocabulary score | -0.017 | 0.251 |
|  | Flanker task score | -0.007 | -3.263** |
|  | Pattern matching score | -0.0005 | -5.824* |
|  | Picture sequence score | -0.043 | -1.406 |
|  | Reading score | 0.002 | -1.033 |
| BRI | Picture vocabulary score | -0.216 | 5.599 |
|  | Flanker task score | -0.212 | -20.209* |
|  | Pattern matching score | -0.192 | -43.202* |
|  | Picture sequence score | -0.597 | -0.249 |
|  | Reading score | -0.068 | -2.785 |
| WHtR | Picture vocabulary score | -3.817 | 75.586 |
|  | Flanker task score | -4.271 | -351.810* |
|  | Pattern matching score | -4.297 | -780.200* |
|  | Picture sequence score | -10.452 | -60.801 |
|  | Reading score | -1.243 | -57.487 |

**Note:** The joint linear mixed-effects models simultaneously evaluate follow-up adiposity and annualized adiposity change ( $\Delta$ ) as independent predictors. $\beta$ -values represent standardized coefficients per SD increase in adiposity metrics. Negative $\Delta\beta$ indicates faster fat accrual associates with poorer cognitive trajectories. Covariates and significance markers consistent with Table 2.

We further examined the longitudinal association between adiposity change and the rate of cognitive development, as indexed by SPC in cognitive task scores over 4-year follow-up. Greater increases in all adiposity indicators were significantly associated with slower improvement in flanker task performance (ΔzBMI: β = −0.005, adjusted p = 0.041; ΔWC: β = −0.001, adjusted p <0.001; ΔBRI: β = −0.006, adjusted p <0.001; ΔWHtR: β = −0.113, adjusted p <0.001), and pattern matching score (ΔzBMI: β = −0.009, adjusted p = 0.041; ΔWC: β = −0.001, adjusted p = 0.008; ΔBRI: β = −0.009, adjusted p = 0.003; ΔWHtR: β = −0.166, adjusted p = 0.003) (Table 4).

**Table 4:** Association between adiposity accrual rate and cognitive development.

| Adiposity | Cognitive task (SPC) | $\Delta (\beta)$ |
| --- | --- | --- |
| zBMI | Picture vocabulary score | -0.003 |
|  | Flanker task score | -0.005* |
|  | Pattern matching score | -0.009* |
|  | Picture sequence score | -0.002 |
|  | Reading score | -0.002 |
| WC | Picture vocabulary score | -0.0001 |
|  | Flanker task score | -0.001*** |
|  | Pattern matching score | -0.001** |
|  | Picture sequence score | -0.001 |
|  | Reading score | -0.0002 |
| BRI | Picture vocabulary score | -0.001 |
|  | Flanker task score | -0.006*** |
|  | Pattern matching score | -0.009 |
|  | Picture sequence score | -0.005 |
|  | Reading score | -0.001 |
| WHtR | Picture vocabulary score | -0.023 |
|  | Flanker task score | -0.113*** |
|  | Pattern matching score | -0.166** |
|  | Picture sequence score | -0.098 |
|  | Reading score | -0.019 |

**Note:** Covariates are identical to Table2, with the addition of baseline adiposity measures and cognitive task scores. Interpretation of $\beta$ -values and significance markers consistent with Table2.

Our longitudinal analyses revealed significant associations between adiposity changes and patterns of cortical development over the 4-year follow-up period (Fig. 3a, Fig.S1). After controlling for covariates, adiposity change was positively associated with SPC in CT of the parietal lobe (e.g., superior parietal lobule: β = 0.001 to 0.093, adjusted p < 0.05; postcentral gyrus: β = 0.001 to 0.090, adjusted p < 0.05) and the right superior temporal sulcus (β = 0.000 to 0.033, adjusted p < 0.05), indicating that the accumulation of adiposity may be associated with slower CT thinning in these regions. ΔBRI and ΔWHtR were significantly associated with SPC in CT of the right superior frontal gyrus (ΔBRI: β = 0.002, adjusted p = 0.020; ΔWHtR: β = 0.044, adjusted p = 0.037) (Fig. 3a). Furthermore, adiposity change was positively associated with SPC in CV of visual brain regions such as the angular gyrus (β = 0.001 to 0.128, adjusted p < 0.05), while ΔzBMI was associated with SPC in CV of the left middle frontal gyrus (β = 0.006, adjusted p = 0.027) and posterior-dorsal part of the cingulate gyrus (β = 0.010, adjusted p = 0.003) et al. (Fig. 3a). In contrast to the baseline analysis (Fig. 2b), ΔzBMI and ΔBRI/WHtR showed very different impacts on longitudinal brain development.

**Figure 3.**
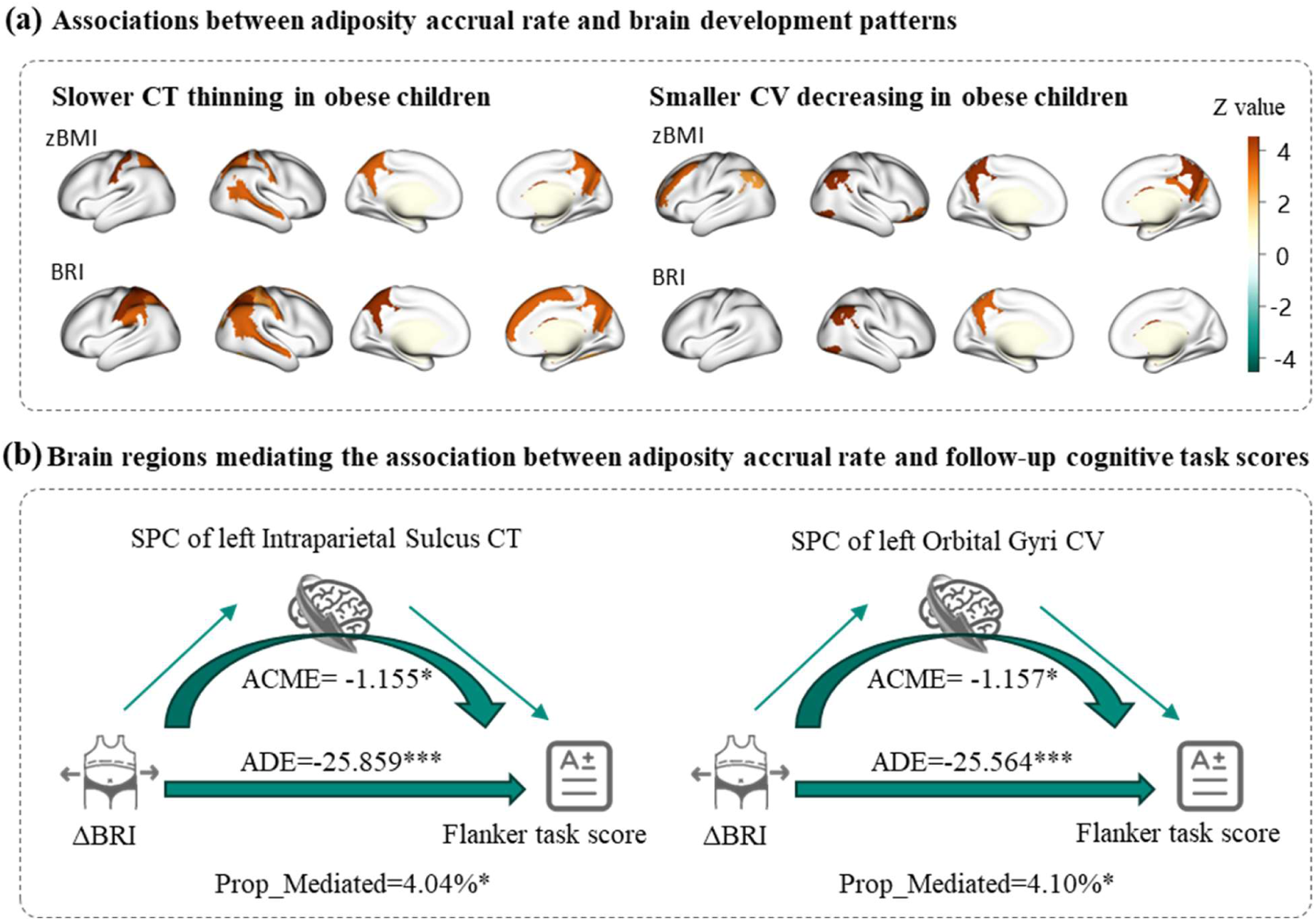
Relations between adiposity accrual rate, neurocognitive characteristics, and brain development. (a) The associations between adiposity accrual rate and SPC of cortical CT or CV. The colored brain areas represented the significant associations after FDR correction, and the value indicated the β-value. (b) Representative mediation models illustrating the indirect effects of ΔBRI on flanker performance via brain development. Abbreviation: BMI, body mass index; BRI, body roundness index; CT, cortical thickness; CV, cortical volume; SPC, symmetrical percentage change; ACME, average casual mediation effect; ADE, average direct effect; Prop_mediated, proportion mediated (ratio of ACME to total effect).

### Longitudinal Mediation by Brain Development in Adiposity-Cognition Relationships

The mediation analyses revealed regionally specific patterns of cortical development mediating the relationship between adiposity changes and cognitive outcomes (Table S3-6). While global mean CT change (SPC) did not show significant mediation effects, several key regions emerged as partial mediators.

For flanker task, left intraparietal sulcus (ACME=-21.22 to −0.12), left postcentral sulcus (ACME=-29.95 to −0.18), and right inferior occipital areas (ACME=-16.62 to −0.88) significantly mediated adiposity change effects.

For reading performance, significant mediating regions were more frequently observed in the right hemisphere, with the posterior lateral sulcus (ACME=-27.34 to −0.18) and intraparietal sulcus (ACME=-25.92 to −0.15) serving as significant mediators. The right medial occipito-temporal and lingual sulci additionally mediated central adiposity effects (ACME=-14.73 to −0.10).

Global cortical volume change (SPC) demonstrated broader mediation effects, significantly explaining relationships between adiposity changes and both flanker task (ACME=-2.17) and reading task performance (ACME=-20.48 to −0.16). Regionally, the left orbital gyri (ACME=-21.29 to −0.15), right angular gyrus (ACME=-27.34 to −0.18), and left inferior frontal gyrus (ACME=-18.32 to −0.13) emerged as most significant mediators for these cognitive domains. In contrast, ΔzBMI showed distinct mediation pathways through left superior temporal sulcus (ACME=-0.1.17), and right intraparietal sulcus (ACME=-1.46) for reading performance. These findings collectively highlight the topographic specificity of how adiposity-related cortical changes may influence different cognitive domains during adolescence.

Collectively, the identified mediators, spanning regions in the intraparietal sulcus, angular gyrus, and inferior frontal cortex, converge on key nodes of the FPN and dorsal attention network (DAN) (Yeo et al., 2011), highlighting the particular vulnerability of higher-order cognitive control and attentional systems to adiposity-related influences during adolescence.

### Central fat trajectories and neurocognitive outcomes in obese adolescents

The subgroup analysis in this section included 730 overweight or obese adolescents (zBMI>=1) with complete follow-up data revealed distinct neurocognitive patterns based on central fat trajectories. The cohort was stratified into two groups: 400 children showing increasing BRI and 330 demonstrating decreasing BRI during follow-up, compared to 1,098 controls maintaining normal zBMI (Table S7, Fig. 4a).

**Figure 4.**
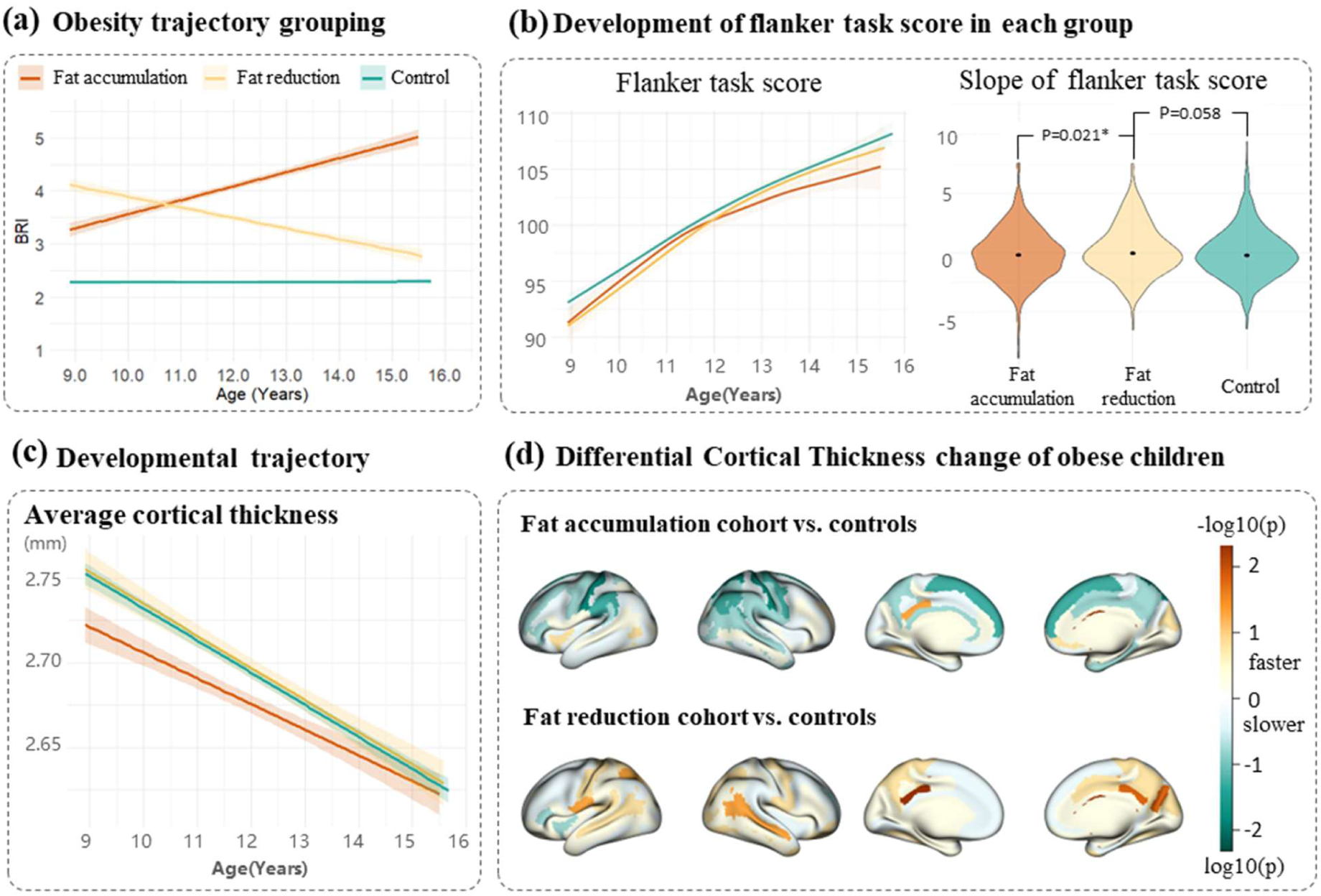
Neurodevelopmental trajectories across central fat change groups. This figure compares brain and cognitive development among obese adolescents with distinct central fat trajectories: increasing (orange), decreasing (yellow), and stable (green) BRI groups. (a) shows group separation in BRI changes over time. (b) reveals differential cognitive changes, with significant group differences in flanker task slopes (*p<0.05, **p<0.01). (c) shows global cortical thickness trajectories across adiposity groups, demonstrating distinct growth pattern among all three groups. (d) presents region specific cortical thickness change patterns in obese versus control groups, with color intensity reflecting statistical significance (−log10[p]).

Cognitive assessments demonstrated divergent outcomes based on adiposity trajectories. Adolescents with decreasing BRI exhibited poor baseline performance (compared to controls: adjusted p=0.024), but significant improvement in the flanker task (compared to controls: adjusted p=0.058; compared to those with increasing BRI: adjusted p = 0.021) (Fig. 4b, Table S8, Table S9).

Adolescents with increasing BRI exhibited an overall pattern of attenuated cortical thinning compared to controls., Given that accelerated cortical thinning during adolescence typically reflects active synaptic pruning and functional specialization(Gogtay et al., 2004), the slowed normative thinning suggests a potential delay in cortical maturation associated. Regionally, in the BRI increase group, slowed CT thinning was most significant in somatosensory areas, including the bilateral postcentral gyrus (p = 0.012-0.020), the right superior parietal lobule (p = 0.027), and the posterior-dorsal cingulate (p = 0.039). Conversely, the decreasing BRI group showed a general trend of accelerated thinning — particularly notable in posterior regions including the parieto-occipital sulcus (p = 0.032) and posterior-dorsal cingulate (p = 0.006-0.029)—which may indicate a catch-up toward typical developmental trajectories following visceral fat reduction (Fig. 4c-d).

CV changes also differed markedly between groups. Adolescents with increasing BRI showed volumetric differences primarily in temporal-occipital regions (inferior occipital gyrus and sulcus p = 0.026), while those with decreasing BRI demonstrated widespread accelerated volume reduction across nearly all cortical regions compared to controls (Fig. S6). These distinct neurocognitive trajectories were replicated in a sensitivity analysis using an alternative classification based on the 25^th^ and 75th percentiles of ΔBRI, confirming the robustness of our findings (Fig. S7).

## Discussion

Our study reveals a complex interplay between adiposity indicators, cognitive outcomes, and neurodevelopment in adolescents. While higher BMI showed selective associations with episodic memory and phonologic processing, central adiposity indices (BRI and WHtR) exhibited broader correlations encompassing inhibitory control and visual processing. These associations were partially mediated by cortical morphology in temporal regions. Compared to static measures at single timepoints, adiposity changes (ΔBRI/ΔWHtR) were shown to be a strong predictor of cognitive performance, particularly for executive functions (flanker task, pattern matching) and memory. Longitudinal analyses revealed that fat accrual rates inversely correlated with cortical thinning rates in frontal-parietal networks. Moreover, the subgroup analyses revealed that the adolescents with reduced central fat exhibited accelerated cortical thinning and volume reduction, a sign of fast brain development; at the same time, this subgroup demonstrated the catch-up of inhibitory control. These findings collectively argue for a paradigm shift in clinical management of childhood obesity: from isolated single adiposity measurements to dynamic monitoring of central fat trajectories during adolescent developmental.

Our cross-sectional findings were consistent with prior research. Previous studies reported that obesity predicts poorer executive function(Ronan et al., 2020) and episodic memory(Sakib et al., 2023), and thinner CT and smaller CV in obese children(Kaltenhauser et al., 2023; Ronan et al., 2020). Extending this work, we demonstrated that central adiposity indices (BRI and WHtR) show broader cognitive associations than BMI, including inhibitory control and visual processing—domains unaffected by generalized adiposity alone. Our analyses also confirmed significant negative associations between adiposity metrics and frontotemporal cortical structure. Moreover, we put these three pieced together and revealed that obesity – cognition relationships were partially mediated by CT and CV in temporal regions, particularly for inhibitory control and episodic memory. This temporal-lobe-centered mediation pattern extends prior studies emphasizing prefrontal mediation(Morys et al., 2024; Ronan et al., 2020), and highlight the need to investigate whether region-specific developmental timing confers differential vulnerability to adiposity-related metabolic stress.

Longitudinal analyses revealed that longitudinal fat accrual velocity outperformed static adiposity measures (baseline or follow-up) in predicting follow-up cognitive outcomes, suggesting that cumulative exposure to central fat may posed greater impact on neurodevelopment. Moreover, accelerated fat accrual was associated with slower frontal and parietal cortical thinning. This deceleration of normative developmental patterns was most pronounced in intraparietal and lingual sulci – regions that functionally support cross-modal information processing (Bogler et al., 2024; Gilmore et al., 2015; Gordon et al., 2018). Mediation analyses also identified these parietal structures as key neural substrates linking fat accrual to poorer follow-up inhibitory control and reading abilities. Notably, longitudinal mediation patterns depended on different types of adiposity indicators. While superior temporal sulcus thinning specifically mediated associations between BMI changes and reading performance, central fat accrual (WC/BRI/WHtR) showed preferential effects through pars triangularis volume reduction. This anatomical dissociation implies differential neural targeting - with central adiposity predominantly impacting prefrontal-limbic circuits involved in cognitive control, whereas generalized adiposity affects posterior cortices supporting perceptual integration.

Furthermore, subgroup analyses revealed that central fat reduction was associated with accelerated cortical thinning and volume loss over four years, particularly in parietal regions, which corresponded to rapid improvements in inhibitory control. This fat-reduction group had lower baseline inhibitory control but achieved catch-up performance during follow-up, suggesting that initial obesity may reflect delayed neurocognitive development rather than irreversible damage and that adverse impact of obesity may resolve with age through neurodevelopmental plasticity. These findings were consistent with previous evidence that a faster rate of cortical thinning during adolescence supports cognitive development, likely reflecting facilitated synaptic pruning (Shaw et al., 2006; Tamnes et al., 2013). In contrast, adolescents with persistent fat accumulation showed slower cortical thinning, highlighting divergent developmental trajectories based on changes in central adiposity. The stratification according to active monitoring of childhood obesity may enable targeted identification of high-risk cohorts requiring neuroprotective management.

Several methodological constraints warrant consideration. First, while BRI and WHtR provide practical anthropometric proxies for central adiposity, they are not direct measurements of compartment-specific fat distribution patterns, which could be more precisely quantified with methods like DXA or MRI. Second, although the 4-year follow-up study represent a substantial observation period, it may not fully capture late-adolescent neurodevelopmental transitions given the extended duration of cortical maturation processes. Third, the absence of serum metabolic biomarkers and fMRI data precludes mechanistic elucidation of the obesity-brain-behavior pathway, particularly regarding neuroinflammatory cascades and reward-processing circuitry engagement. Finally, potential confounders including heterogeneous sleep patterns and pharmaceutical interventions may influence the observed effect.

## Data Availability

Data used in the preparation of this article were obtained from the Adolescent Brain Cognitive DevelopmentSM (ABCD) Study (https://abcdstudy.org), held in the NIMH Data Archive (NDA). This is a multisite, longitudinal study designed to recruit more than 10,000 children age 9-10 and follow them over 10 years into early adulthood.

## Acknowledgement

This work was supported by the Ministry of Science and Technology of the People’s Republic of China (grant no. 2021ZD0200202), the National Natural Science Foundation of China (grant nos. 32427802 and U24A20754). Data used in the preparation of this article were obtained from the Adolescent Brain Cognitive Development SM (ABCD) Study (https://abcdstudy.org), held in the NIMH Data Archive (NDA). This is a multisite, longitudinal study designed to recruit more than 10,000 children age 9-10 and follow them over 10 years into early adulthood. The ABCD Study® is supported by the National Institutes of Health and additional federal partners under award numbers U01DA041048, U01DA050989, U01DA051016, U01DA041022, U01DA051018, U01DA051037, U01DA050987, U01DA041174, U01DA041106, U01DA041117, U01DA041028, U01DA041134, U01DA050988, U01DA051039, U01DA041156, U01DA041025, U01DA041120, U01DA051038, U01DA041148, U01DA041093, U01DA041089, U24DA041123, U24DA041147. A full list of supporters is available at https://abcdstudy.org/federal-partners.html. A listing of participating sites and a complete listing of the study investigators can be found at https://abcdstudy.org/consortium_members/. ABCD consortium investigators designed and implemented the study and/or provided data but did not necessarily participate in the analysis or writing of this report. This manuscript reflects the views of the authors and may not reflect the opinions or views of the NIH or ABCD consortium investigators.

